# Empagliflozin Improves Mitochondrial Biogenesis and Ameliorates Experimental Pulmonary Vascular Remodeling, But May Not Benefit Patients with Pulmonary Arterial Hypertension

**DOI:** 10.1101/2025.01.27.25321226

**Authors:** Keimei Yoshida, Eszter Toth, Erik Duijvelaar, Beau Neep, Stuti Agarwal, Xiaoke Pan, Takayuki Sanada, Yu Yoshida, Jurjan Aman, Vinicio A. de Jesus Perez, M. Louis Handoko, Frances S. de Man, Xiao-Qing Sun, Harm-Jan Bogaard

**Affiliations:** Department of Cardiovascular Medicine, Faculty of Medical Sciences, Kyushu University, Fukuoka, Japan; Department of Pulmonary Medicine, Amsterdam UMC, Vrije Universiteit, Amsterdam, The Netherlands; Division of Pulmonary, Allergy, and Critical Care Medicine, Wall Center for Cardiopulmonary Research, Stanford University, Stanford, CA, United States; Department of Cardiology, University Medical Center Utrecht, The Netherlands

**Author notes:** **Corresponding:** Keimei Yoshida, MD, PhD, Kyushu University, 3-1-1, Maidashi, Higashiku, Fukuoka, Japan. E mail. Eszter Toth and Erik Duijvelaar contributed equally. Xiao-Qing Sun and Harm-Jan Bogaard contributed equally.

## Abstract

**Background:** The sodium glucose cotransporter 2 (SGLT2) inhibitor may improve mitochondrial biogenesis and attenuate pulmonary vascular remodeling in pulmonary arterial hypertension (PAH). We investigated the impact of empagliflozin in PAH.

**Methods:** Lung sections and primary cell cultures isolated from microvascular endothelial cells (MVECs) were collected from control subjects and PAH patients. MVECs were treated with empagliflozin and mitochondrial biogenesis, cell metabolism, oxidative stress and cell proliferation were evaluated. Subsequently, PAH was induced in male and female rats (n=12 respectively) with SU5416 injection (25 mg/kg s.c.) followed by 3 weeks of hypoxia (10% O_2_), the experimental PAH model known to mimic human PAH pathophysiology. Four weeks after SU5416 injection, rats were treated by empagliflozin (300 mg/kg chow, n=12) or placebo (n=12) for 4 weeks and hemodynamic, protein and histological analyses were performed. In addition, we conducted a phase IIa proof of concept trial, EMPHOWER, to assess the feasibility of 12 weeks of empagliflozin treatment in PAH patients.

**Results:** Immunofluorescent staining of human lung tissue showed expression of SGLT2 in the intima of small pulmonary arteries from PAH patients, not controls. In comparison to control MVECs, PAH MVECs showed increased protein expression of SGLT2, along with decreased expression of the peroxisome proliferator-activated receptor gamma coactivator-1α. Furthermore, empagliflozin enhanced expression of mitochondrial encoded genes and mitochondrial respiration, suggesting increased mitochondrial biogenesis. Moreover, empagliflozin significantly attenuated oxidative stress and proliferation of PAH MVECs. In SuHx rats, chronic treatment with empagliflozin significantly reduced pulmonary vascular resistance and thickening of the intima of small pulmonary arteries. Finally, 8 patients diagnosed with idiopathic and heritable PAH were enrolled in the phase IIa EMPHOWER trial. There was no discontinuation of empagliflozin during the study period and there were no treatment associated serious adverse events. There were no changes in biomarkers, WHO functional class, six-minute walk distance, or EMPHASIS score. However, RV ejection fraction and RV global longitudinal strain slightly worsened after empagliflozin treatment (from 45 ± 10% to 38 ± 12%, P=0.036, and from −15.2 ± 4.2% to −13.2 ± 3.96%, P=0.002, respectively).

**Conclusion:** SGLT2 expression is increased in the PAH endothelium. Treatment with empagliflozin improves mitochondrial biogenesis and attenuates proliferation of PAH MVECs. Empagliflozin attenuates pulmonary vascular remodeling in experimental PAH. While twelve weeks of empagliflozin treatment seemed feasible in patients with idiopathic or hereditary PAH, we observed signs of RV deterioration.

**Clinical perspective:** **What is new?**

- This is the first study to demonstrate the role of sodium glucose cotransporter 2 (SGLT2) in the endothelial cell proliferation of pulmonary arterial hypertension (PAH).
- SGLT2 was expressed and increased in microvascular endothelial cells (MVECs) from the lung of PAH patients accompanied by suppression of peroxisome proliferator-activated receptor gamma coactivator-1α.
- Empagliflozin improved mitochondrial biogenesis and respiration in PAH MVECs.
- Empagliflozin reversed pulmonary angioproliferation and attenuated pulmonary vascular resistance in experimental PAH rats.
- The EMPHOWER PoC study showed the feasibility of 12 weeks of empagliflozin treatment in PAH, however, right ventricular function assessed by cardiac magnetic resonance imaging worsened.

**What are the clinical implications**

- SGLT2 inhibition may improve mitochondrial respiration and reverse pulmonary vascular remodeling in PAH.
- The effect of empagliflozin on right ventricular function requires further caution and investigation.

## Introduction

Pulmonary arterial hypertension (PAH) is a devastating disease characterized by progressive pulmonary angioproliferation and increased pulmonary vascular resistance (PVR).^1^ After an initial period of increased right ventricular (RV) contractility to compensate for the increased afterload, cardiac output (CO) eventually decreases, ultimately leading to RV failure and premature death.^2^ Despite recent advances in the treatment of PAH, its prognosis remains poor.^3^

Recently, the anti-diabetic drug empagliflozin, a sodium glucose cotransporter 2 (SGLT2) inhibitor, was shown to improve the outcome of patients with heart failure, regardless of etiology and irrespective of the presence of diabetes mellitus.^4,5^ Interestingly, apart from expected effects such as glucosuria and natriuresis, empagliflozin was shown to have pleiotropic effects, including mitigation of oxidative stress, inflammation and sympathoexcitation.^6^ Moreover, several preclinical studies have suggested that these pleiotropic effects are also present in endothelial cells: critical cells in many cardiovascular conditions that were initially thought not to express SGLT2 (gene name: *SLC5A2*). Yet, SGLT2 inhibitors ameliorate endothelial dysfunction through a reduction of reactive oxygen species (ROS) generation,^7^ mitigation of inflammatory responses,^8^ and suppressed expression of adhesion molecules.^9^ These effects were demonstrated in human coronary artery and aortic endothelial cells.^10^ Importantly, the effects of SGLT2 inhibitors on the pulmonary vasculature remain obscure. Although one preclinical study suggested a beneficial effect of SGLT2 inhibition in rats with experimental pulmonary hypertension (PH),^11^ the study lacked mechanistic data to answer the question whether this benefit was mediated through improved endothelial function. In particular, the role of SGLT2 in pulmonary macro-or microvascular endothelial cells has not been studied before.

Metabolic abnormalities play an important role in the pathogenesis of PAH.^12^ Mitochondrial dysfunction and excessive glycolysis cause a phenotypic switch of PAH MVECs, characterized by hyperproliferation and apoptosis resistance.^13^ Peroxisome proliferator-activated receptor gamma coactivator-1α (PGC-1α), a transcriptional coactivator that regulates energy metabolism through stimulation of mitochondrial biogenesis,^14^ has been implicated in PH development in both animal models and human studies. An animal study showed that enhancing expression of PGC-1α attenuates hypoxia-induced PH in mice,^15^ and a clinical study showed that PGC-1α inversely correlates with disease severity in patients with idiopathic PAH.^16^ However, its interaction with SGLT2 and its role in PAH MVECs remains undetermined.

Here, we hypothesized that empagliflozin improves mitochondrial biogenesis and reverses pulmonary vascular remodeling in an experimental PAH. We demonstrate that empagliflozin enhances PGC-1α expression, improves mitochondrial biogenesis and respiration, and restores dysfunction of MVECs derived from PAH patients. Moreover, we show that empagliflozin reverses pulmonary vascular remodeling and PH in a rat model of severe angioproliferative PAH. Lastly, we performed a phase IIa proof of concept (PoC) clinical trial to test the feasibility of 12 weeks of empagliflozin treatment in idiopathic and heritable PAH patients (ClinicalTrials.gov number, NCT05493371).

## Methods

### Empagliflozin preclinical study

#### Primary cell isolation

Lobectomy tissue was used to isolate control (CTRL) MVECs, while PAH MVECs were isolated from ex-planted lung tissue from patients diagnosed with idiopathic PAH. Details on endothelial cell isolation and culture were published before.^17^ The study was approved by the institutional review of board of the Amsterdam University Medical Center (Amsterdam, the Netherlands) and informed consent was given.

After starvation for 4 hours with 1% Fetal Bovine Serum (FBS), MVECs were treated for 1 hour by either 1 μM empagliflozin (HY-15409, MedChem Express, Monmouth Junction, NJ, USA) dissolved in dimethyl sulfoxide (DMSO) or DMSO without empagliflozin. Lysates were collected for each experimental condition.

#### siRNA and plasmid transfection

Knockdown was performed as described previously.^18^ MVECs were grown until 60% confluence and medium was refreshed 24 hours before transfection. To knockdown the protein of interest, ON-TARGETplus human siRNA against SLC5A2 (L-007590-00-0005), SIRT1 (L-003540-00-0005) and non-targeting control (NT, D-001810-10-05) were purchased from Dharmacon. To overexpress SGLT2, plasmid coding SLC5A2 (#132241) was purchased from Addgene. After bacteria were selected by broth containing Kanamycin, plasmid constructs were collected. MVECs were transfected with a final concentration of 25 nM siRNA or 5 nM plasmid construct using DharmeFECT1 (T2002-03, Dharmacon, USA). Transfection was performed overnight with 10% FBS in ECM. Medium was refreshed with cECM the next day. Cells were used for experiments 48 hours after transfection.

#### MitoTracker analysis

To detect mitochondrial reactive oxygen species, we performed fluorescent analysis using MitoTracker Red CM-H_2_Xros (M7513, Invitrogen).^7^ The Procedure was performed following the description in the data sheet. Quantification of fluorescence signal was performed by SpectraMax iD3 (Molecular Devices, UK).

#### Seahorse analysis

Seahorse analysis was performed to evaluate the cellular metabolism of MVECs as described previously.^19^ Mitochondrial respiration and aerobic glycolysis were monitored in real time with the Seahorse Extracellular Flux Analyzer (XF96; Agilen, USA) by measuring the oxygen consumption rate (OCR; indicative of respiration) and extracellular acidification rate (ECAR; indicative of glycolysis) using the Mitochondrial Stress Test assay and the Glycolytic Rate assay. Values were normalized to the total protein concentration per well assessed by BCA assay (Thermofisher Scientific, USA) after completion of the XF assay.

#### Empagliflozin in experimental pulmonary arterial hypertension

(Figure S1, n=30) The study was approved by the independent local animal ethics committee at Amsterdam University Medical Center (study number: 9866-Long21-05) and was performed in compliance with the guidelines issued by the Dutch government.

Male and female Sprague-Dawley rats (n=15 in each sex, Envigo) were used throughout the experiments. Rats were housed in standard conditions and food and water was available *ad libitum*. PH was induced by SU5416 and hypoxia (SuHx) according to previous reports.^20,21^ In brief, rats were subjected to a single subcutaneous injection of vascular endothelial growth factor inhibitor SU5416 (25 mg/kg, Tocris Bioscience) followed by a 3-week exposure to 10% hypoxia, after which they returned to normoxia. After demonstration of the presence of PH at week 4 using echocardiography, animals were randomized into two groups, receiving empagliflozin (male/female =6/6) or placebo (male/female =6/6). Rats were treated for 4 weeks with empagliflozin mixed chow (300 mg/kg) or placebo chow, provided by Boehringer Ingelheim (Pharma GmbH & Co. KG, Germany). At week 8, we performed echocardiography and right heart catheterization (supplemental methods), while organs were harvested after euthanizing the animals. We also included control rats (n=3 for each sex) in the study, treated with the same chow as the placebo group.

#### Histopathological analysis of the pulmonary circulation

To determine the severity of pulmonary vascular remodeling, we performed Elastica van Gieson (EvG) stainings and quantified the ratio of occlusive lesions, as well as intima and media wall thickness, as previously described.^22^ A minimum of thirty randomly distributed and transversely cut pulmonary arterioles, cut with an outer diameter between 25 and 100 μm, were measured.

### Empagliflozin clinical study

EMPHOWER was a single-center, open-label, single-arm interventional PoC study. Given the exploratory design of the study, no formal sample size calculation was performed. Approval of the trial protocol was granted by the institutional review board of the Amsterdam UMC, location VUmc. This study was registered with the EU Clinical Trials Register (EudraCT 2022-002400-20) and Clinicaltrials.gov (NCT05493371). See supplement for the complete list of inclusion and exclusion criteria. The primary endpoint comprised the tolerability, feasibility and safety of 12 weeks of treatment with 10mg of oral empagliflozin once daily as add-on therapy. Tolerability was assessed by determining the number of patients who had to prematurely discontinue treatment due to intolerability or adverse events. Feasibility was comprised the time needed to include a total of 8 patients. Secondary endpoints included assessments of right heart function using cardiac magnetic resonance imaging (CMR) and transthoracic echocardiography (TTE), blood biomarkers, functional class, 6-minute walking distance (6MWD) and quality of life. Given the single-arm design, measurements taken before first drug administration were compared with those obtained after 12 weeks of treatment within the same patient.

### Statistical Analysis

Statistical analyses were performed using GraphPad Prism 9 (GraphPad Software, San Diego, CA, USA) and R version 4.1.2. In the preclinical study, paired or unpaired student’s t-tests were used for comparisons between two groups. Multiple comparisons were assessed by one-way analysis of variance, followed by post-hoc Tukey-Kramer test. Two-way analysis of variance followed by post-hoc Tukey-Kramer test was used for repeated data of echocardiography analysis and morphological analysis. Data are expressed as mean ± SEM. In the clinical study, no statistical analyses were performed on the primary endpoints. Other endpoints with two continuous outcomes were analyzed using a paired sample t-test or linear mixed models when more than two time points were available. Endpoints with a categorical or ordinal outcome were analyzed using the chi-squared test or Wilcoxon signed rank test respectively. Differences were considered significant when p < 0.05.

## Results

### Increased expression of SGLT2 in PAH MVECs

Figure 1A shows immunofluorescent staining of SGLT2 in lung tissue from PAH patients, while staining is absent in control subjects (CTRL). In PAH, SGLT2 is present in the intima, as shown by co-localization with von Willebrand Factor (vWF). Figure 1B shows immunofluorescent staining of SGLT2 in MVECs from CTRL and PAH. While SGLT2 is present in cells from control subjects, the staining is only nuclear and less intense than in PAH MVECs, in which cells SGLT2 is also present in the cytoplasm. Moreover, PAH MVECs show broader alignment of VE-Cadherin and co-localization of SGLT2 and VE-cadherin. Immunoblotting confirmed higher expression of SGLT2 in PAH MVECs compared to CTRL (Figure 1C and 1D). On the other hand, the expression of PGC-1α was significantly decreased in PAH MVECs (Figure 1C and 1E). We observed an inverse relationship between SGLT2 and PGC-1α (Figure 1F). These data suggest decreased mitochondrial biogenesis in the pulmonary vasculature of PAH.

**Figure 1.**
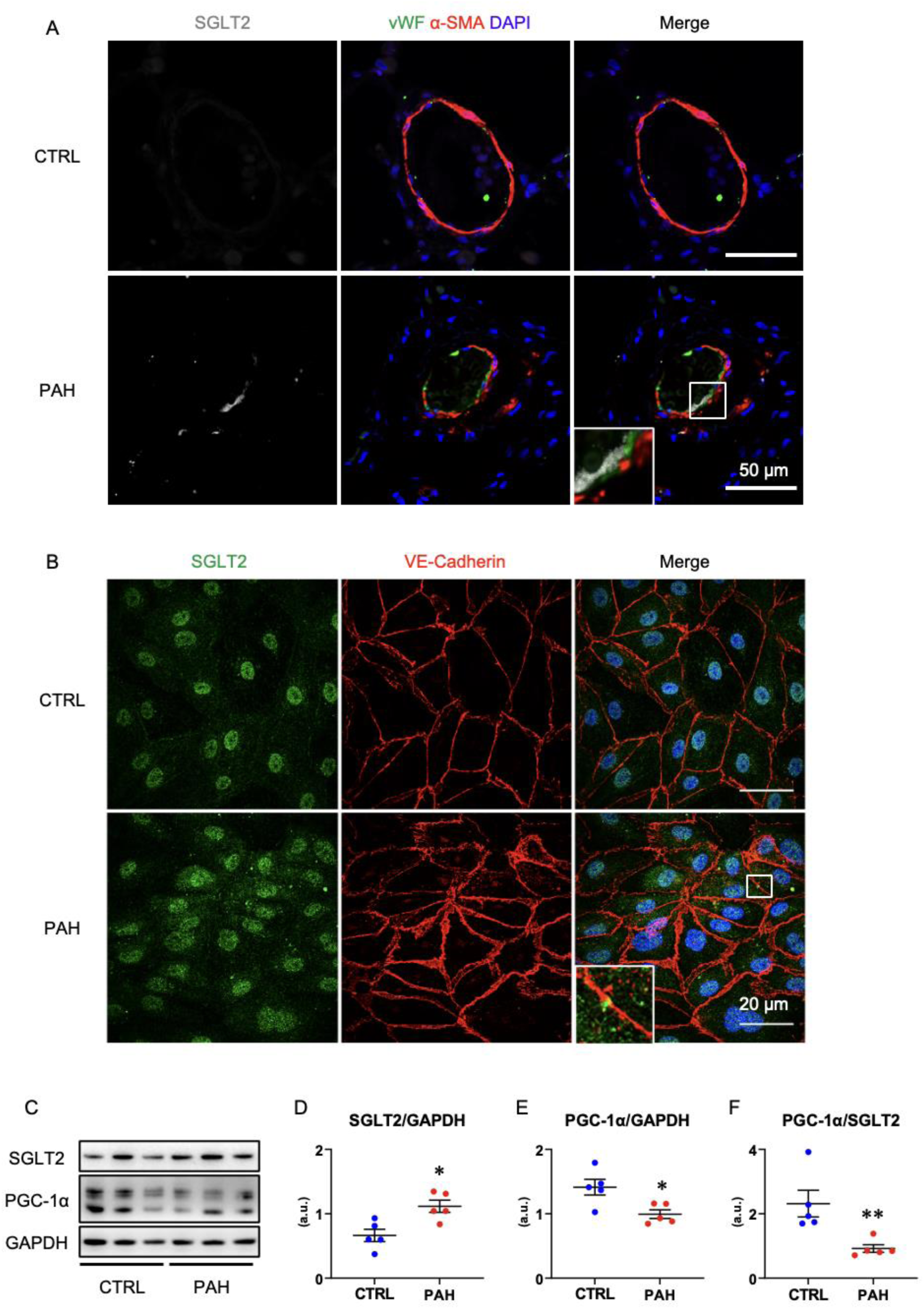
Expression of SGLT2 in pulmonary artery and MVECs. **A**, Representative immunofluorescent staining of SGLT2 (Gray), von Willebrand Factor (vWF, Green), and DAPI (Blue) of pulmonary artery from CTRL and PAH. **B**, Representative immunofluorescent staining of SGLT2 (Green), VE-Cadherin (Red), and DAPI (Blue) of MVECs from CTRL and PAH. **C** through **F**, Representative western blot and quantification of SGLT2 and PGC-1α of MVECs from CTRL and PAH, normalized by GAPDH. Values are mean ± SEM. Difference was tested by using Mann-Whitney test. *p<0.05, **p<0.01.

### SGLT2 inhibition by empagliflozin enhances expression of PGC-1α via SIRT1 in PAH MVECs

We next evaluated the effect of empagliflozin on PAH MVECs. Empagliflozin treatment significantly increased the expression of PGC-1α as well as SGLT2 (Figure 2A, 2B and 2C). To evaluate whether the effect of empagliflozin on PGC-1α was mediated by inhibition of SGLT2, we performed knockdown of SGLT2 (gene name: *SLC5A2*) in MVECs using siRNA. In the SLC5A2 siRNA transfected MVECs, expression of SGLT2 was decreased while PGC-1α was increased (Figure 2D, 2E and 2F). Moreover, overexpression of SGLT2 resulted in downregulation of PGC-1α (Figure 2G, 2H and 2I). These data suggest that SGLT2 negatively regulates the expression of PGC-1α in MVECs.

**Figure 2.**
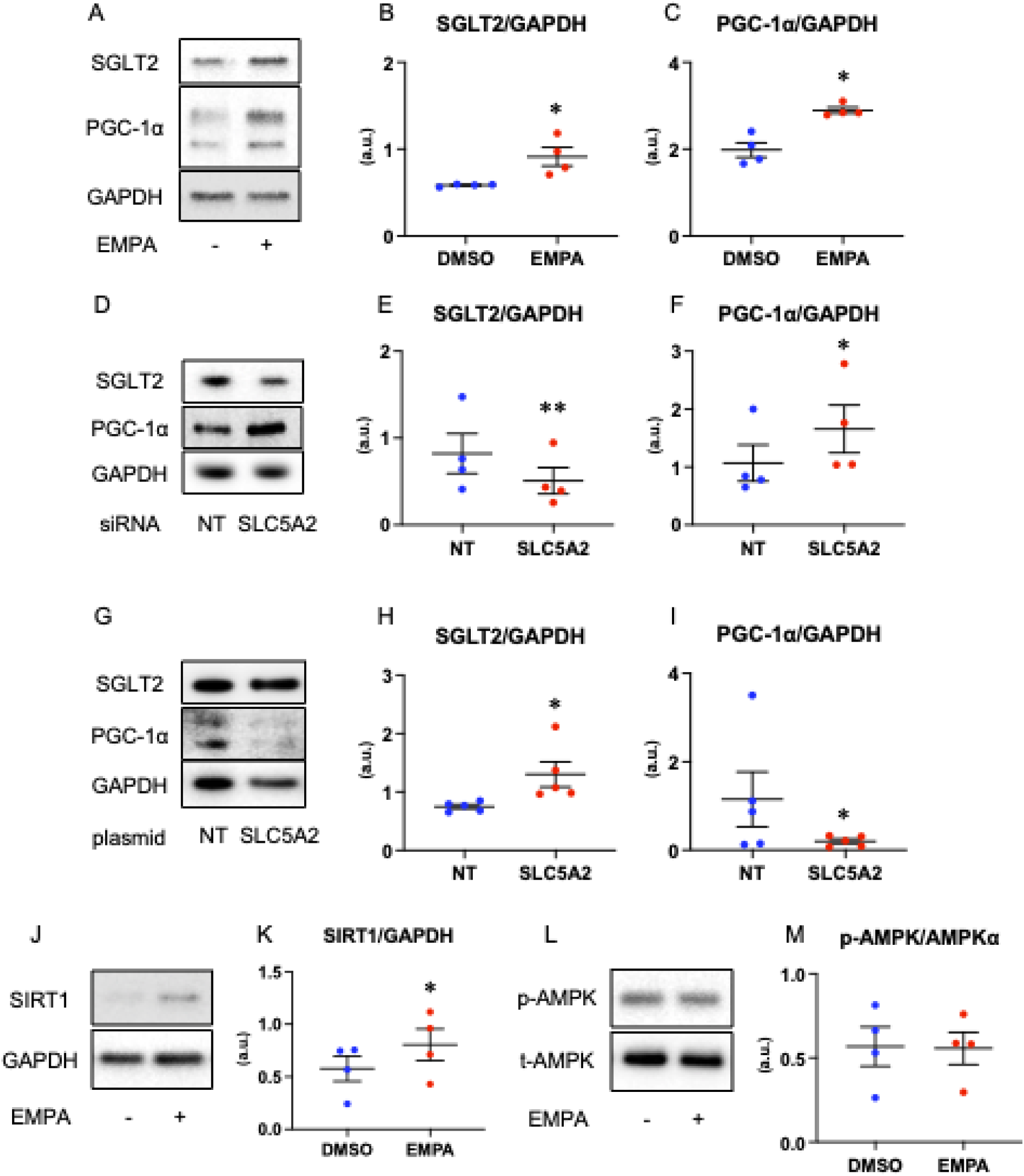
The effect of empagliflozin and SGLT2 modulation on PAH MVECs. **A** through **C**, Representative western blot and quantification of SGLT2 and PGC-1α of PAH MVECs treated with empagliflozin (EMPA) or DMSO. **D** and **E**, Representative western blot and quantification of SIRT1 of PAH MVECs treated with EMPA or DMSO, normalized by GAPDH. **F** and **G**, Representative western blot and quantification of p-AMPK of PAH MVECs treated with EMPA or DMSO, normalized by t-AMPK. **H** through **I**, Representative western blot and quantification of BMPR2, p-Smad1/5/9 and p-Smad2/3 of PAH MVECs treated with EMPA or DMSO, normalized by GAPDH. **J** and **K**, Representative western blot and quantification of SIRT1 of PAH MVECs treated with EMPA or DMSO, normalized by GAPDH. **L** and **M**, Representative western blot and quantification of p-AMPK of PAH MVECs treated with EMPA or DMSO, normalized by t-AMPK. Values are mean ± SEM. Difference was tested by using Paired t-test. *p<0.05, **p<0.01.

Because PGC-1α is regulated by sirtuin1 (SIRT1) and phosphorylated AMP-activated protein kinase (p-AMPK), we next assessed the effect of empagliflozin on their expression. The expression of SIRT1 was significantly increased after empagliflozin treatment (Figure 2J and 2K) while there was no difference in p-AMPK (Figure 2L and 2M). This data suggests that empagliflozin treatment enhanced the expression of PGC-1α possibly via SIRT1.

### SGLT2 inhibition by empagliflozin increases mitochondrial biogenesis and respiration, suppresses oxidative stress and cell proliferation in PAH MVEC

Next, we investigated the role of SGLT2 in mitochondrial biogenesis. Empagliflozin treatment enhanced expression of mitochondria encoded genes (Figure 3A, 3B and 3C). These data further support that empagliflozin enhances mitochondrial biogenesis in PAH MVECs. Furthermore, we performed Seahorse analysis to evaluate the effect of empagliflozin on metabolism in MVECs. Empagliflozin significantly increased OCR and reduced ECAR in PAH MVECs (Figure 3D and 3E). These data suggest that empagliflozin enhances mitochondrial respiration in PAH MVECs.

**Figure 3.**
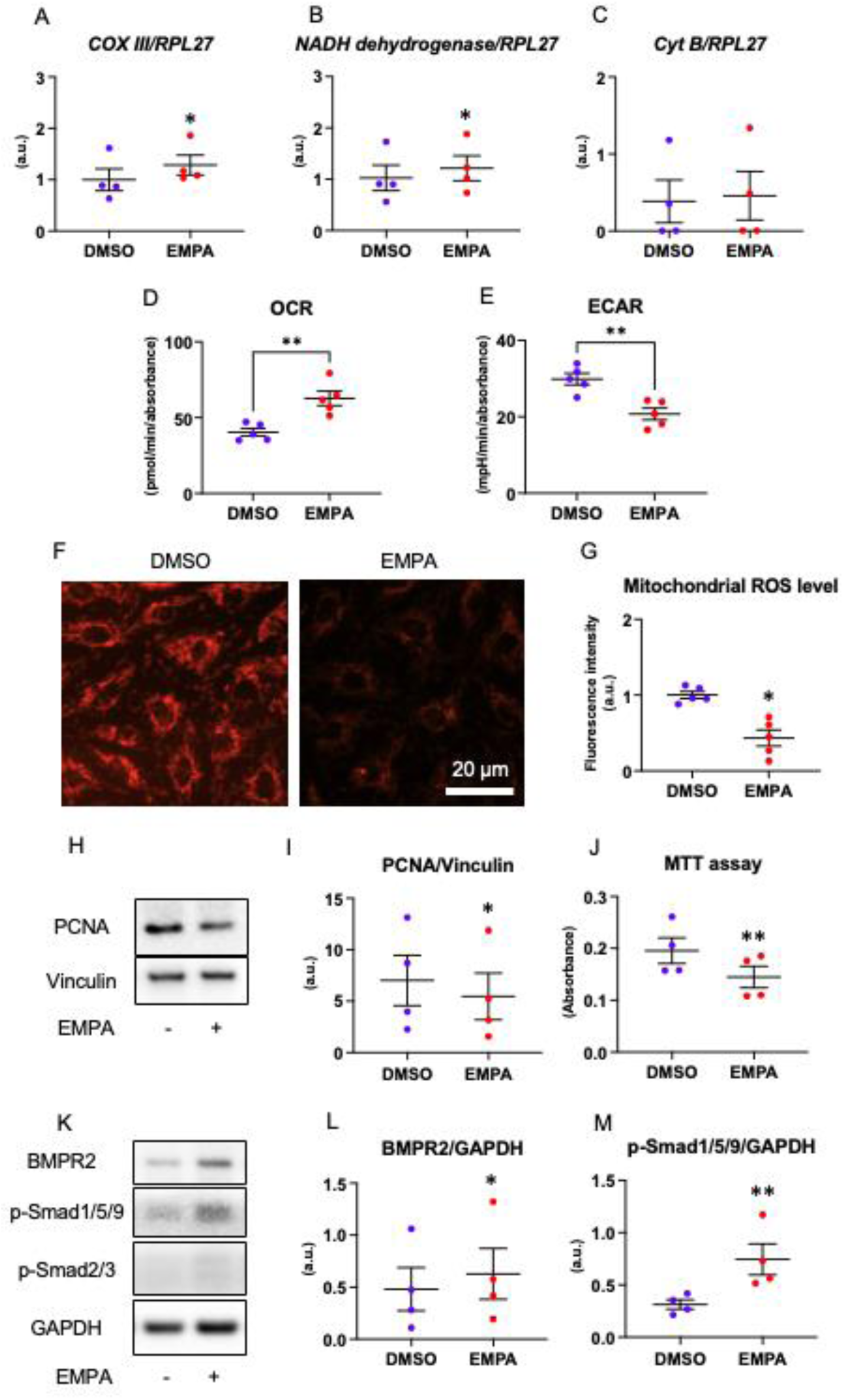
The effect of empagliflozin treatment on metabolism and oxidative stress in PAH MVECs. **A** through **C**, Quantification of gene level of COXIII, NADH dehydrogenase and Cyt B of PAH MVECs treated by EMPA or DMSO, normalized by RLP27. **D** and **E**, Oxygen consumption rate (OCR) and extracellular acidification rate (ECAR) of PAH MVECs treated by EMPA or DMSO. **F** and **G**, Representative image of using MitoTracker to detect the mitochondrial reactive oxygen species (ROS) and quantification of PAH MVECs treated with empagliflozin (EMPA) or DMSO. **H** and **I**, Representative western blot and quantification of PCNA of PAH MVECs treated with EMPA or DMSO, normalized by Vinculin. **J,** Evaluation of cell proliferation using MTT assay in PAH MVECs. **K** through **M**, Representative western blot and quantification of BMPR2, p-Smad1/5/9 and p-Smad2/3 of PAH MVECs treated with EMPA or DMSO, normalized by GAPDH. Values are mean ± SEM. Difference was tested by using Paired t-test. *p<0.05, **p<0.01.

As oxidative stress is deeply related with mitochondrial function and biogenesis, we evaluated mitochondrial ROS in MVECs. Fluorescent live-cell imaging using MitoTracker showed that the expression of mitochondrial ROS was suppressed in empagliflozin treated MVECs compared to MVECs treated with DMSO (Figure 3F). Fluorescent analysis showed significant reduction of the mitochondrial ROS level in empagliflozin treated PAH MVECs (Figure 3G).

We also evaluated the effect of empagliflozin on cell proliferation. Immunoblotting showed decreased expression of proliferating cell nuclear antigen (PCNA) in empagliflozin treated MVECs (Figure 3H and 3I). In addition, quantitative cell proliferation evaluation with MTT assay showed that empagliflozin treatment significantly attenuated proliferation of PAH MVECs (Figure 3J). Meanwhile empagliflozin did not affect cell proliferation in CTRL MVECs (Figure S2).

Moreover, we evaluated bone morphogenetic protein receptor type-2 (BMPR2)-Smad1/5/9 signaling, as this signaling pathway is known to play an important role in pulmonary vascular homeostasis and is critically involved in the development of PAH. Immunoblotting showed that empagliflozin treatment significantly increased the expression of BMPR2 and phospho-Smad1/5/9 in PAH MVECs (Figure 3K, 3L and 3M).

### Chronic empagliflozin treatment reduces pulmonary vascular resistance, preserves right ventricular function and reduces right ventricular hypertrophy in an experimental PAH model

Next, we evaluated the treatment effect of empagliflozin in SuHx rats. Rats were treated for 4 weeks with empagliflozin mixed chow (300 mg/kg) or placebo chow, initiated 4 weeks after the SU5416 injection (Figure S1). There was no significant difference of body weight progression between empagliflozin and placebo in each sex (Figure S3A). Compared to placebo, rats treated with empagliflozin increased consumption of food and water until the end of the experiment (Figure S3B and S3C). Empagliflozin treated rats were estimated to ingest empagliflozin in the range of 20 to 30 mg/kg/day (Figure S3D).

There was no significant difference in blood glucose levels among groups (Figure S4A). Of note, there was no hypoglycemia in the empagliflozin group. Empagliflozin significantly induced glucosuria and urine ketone concentration (Figure S4B and S4C). Empagliflozin decreased urine protein compared to placebo (Figure S4D).

Figure 4A through 4G shows the results of echocardiography at two time points, before and after empagliflozin treatment. At Week 4, there were no differences in any of the measured parameters between placebo and empagliflozin. At Week 8, empagliflozin treated rats showed significantly decreased RV dimensions, attenuated RVWT, increased TAPSEI and increased CI. Empagliflozin also attenuated eRVSP and eTPRI. These data suggest that empagliflozin decreases RV afterload and improves RV function in SuHx rats.

**Figure 4.**
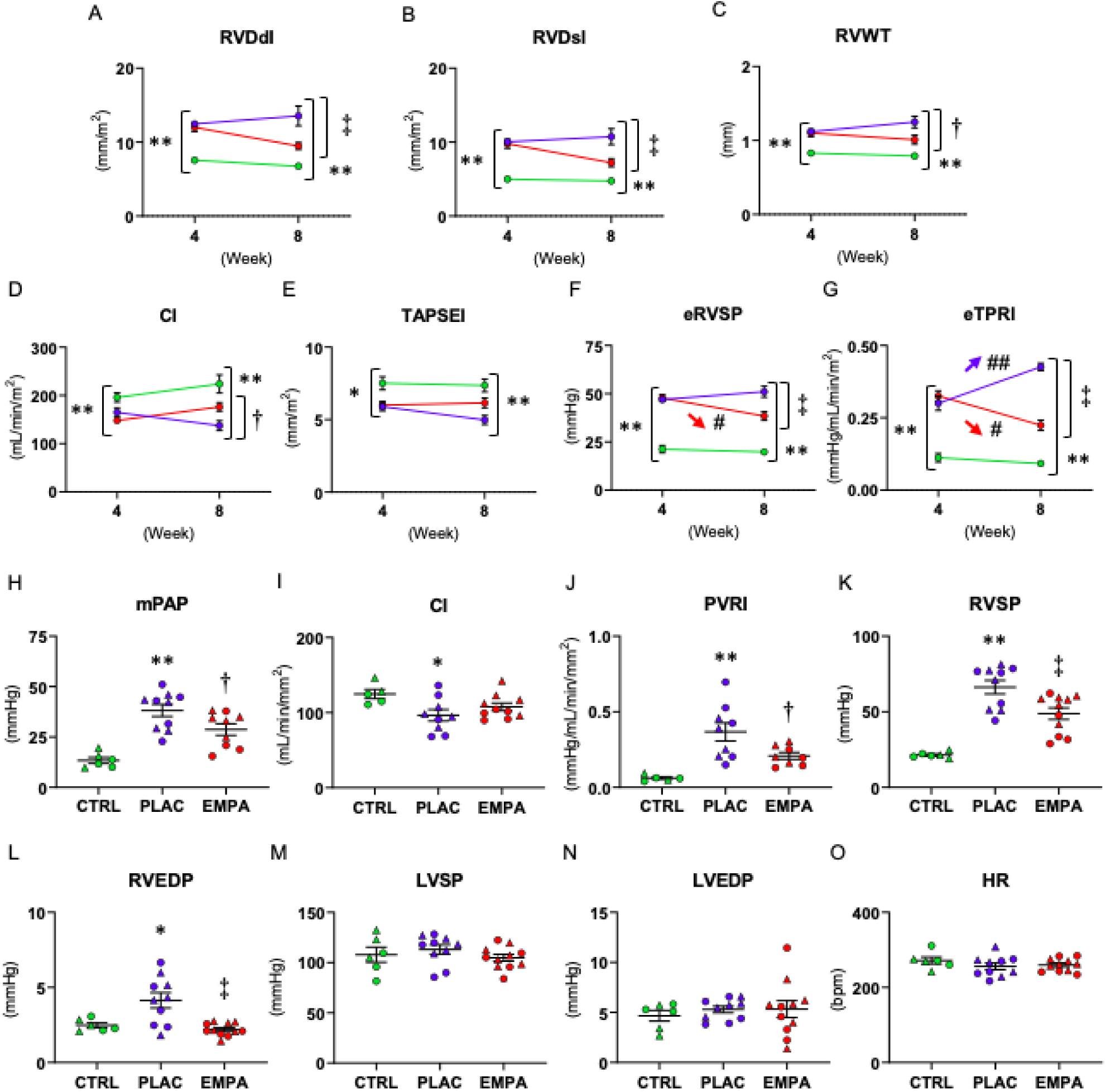
Empagliflozin reduces pulmonary vascular resistance and preserves RV function. The results of echocardiography from control (CTRL, green), and placebo (PLAC, blue) or empagliflozin (EMPA, red) treated PAH rats in 2 time points. Right ventricular dimension diastole index (RVDdI, **A**), right ventricular dimension systole index (RVDsI, **B**), right ventricular wall thickness (RVWT, **C**), cardiac index (CI, **D**), tricuspid annulus plane systolic excursion index (TAPSEI, **E**), estimated right ventricular systolic pressure (eRVSP, **F**), and estimated total pulmonary resistance index (eTPRI, **G**). Values are mean ± SEM. Difference was tested by using two-way analysis of variance followed by post-hoc Tukey’s multiple comparisons test. ** p<0.01, * p<0.05 vs. CTRL, ‡ p<0.01, † p<0.05 vs. PLAC, ## p<0.01, # p<0.05 vs. Week 4. The results of catheterization from control (CTRL), and placebo (PLAC) or empagliflozin (EMPA) treated PAH rats (Female; circle, Male; triangle). Mean pulmonary artery pressure (mPAP, **H**), cardiac index (CI, **I**), pulmonary vascular resistance index (PVRI, **J**), right ventricular systolic pressure (RVSP, **K**), right ventricular end-diastolic pressure (RVEDP, **L**), left ventricular systolic pressure (LVSP, **M**), left ventricular end-diastolic pressure (LVEDP, **N**), and heart rate (HR, **O**). Values are mean ± SEM. Difference was tested by using one-way analysis of variance followed by post-hoc Tukey’s multiple comparisons test. ** p<0.01, * p<0.05 vs. CTRL, ‡ p<0.01, † p<0.05 vs. PLAC.

Figure 4H through 4O shows the results of the hemodynamic analyses. Chronic treatment with empagliflozin in PAH rats significantly reduced mPAP, RVSP and PVRI compared to placebo. In placebo, CI was significantly decreased but not in empagliflozin treated rats. Moreover, empagliflozin treatment significantly reduced RVEDP. There was no difference in systemic blood pressure or in the first derivation of RV pressure (Figure S5). Morphological analysis showed a reduction in Fulton index with empagliflozin treatment (Table S1). These data indicate that empagliflozin treatment reduces pulmonary vascular resistance and RV afterload and prevents worsening of RV function in SuHx rats.

### Chronic empagliflozin treatment enhances expression of PGC-1α and suppresses oxidative stress concomitant with reduced pulmonary vascular occlusive lesions

EvG staining showed that empagliflozin treatment lowered the proportion of pulmonary occlusive lesions compared to placebo (Figure 5A and 5B). Empagliflozin also attenuated intima and medial thickness (Figure 5C and 5D). These data suggest that empagliflozin reduces pulmonary vascular resistance via amelioration of pulmonary vascular remodeling, especially intima thickness.

**Figure 5.**
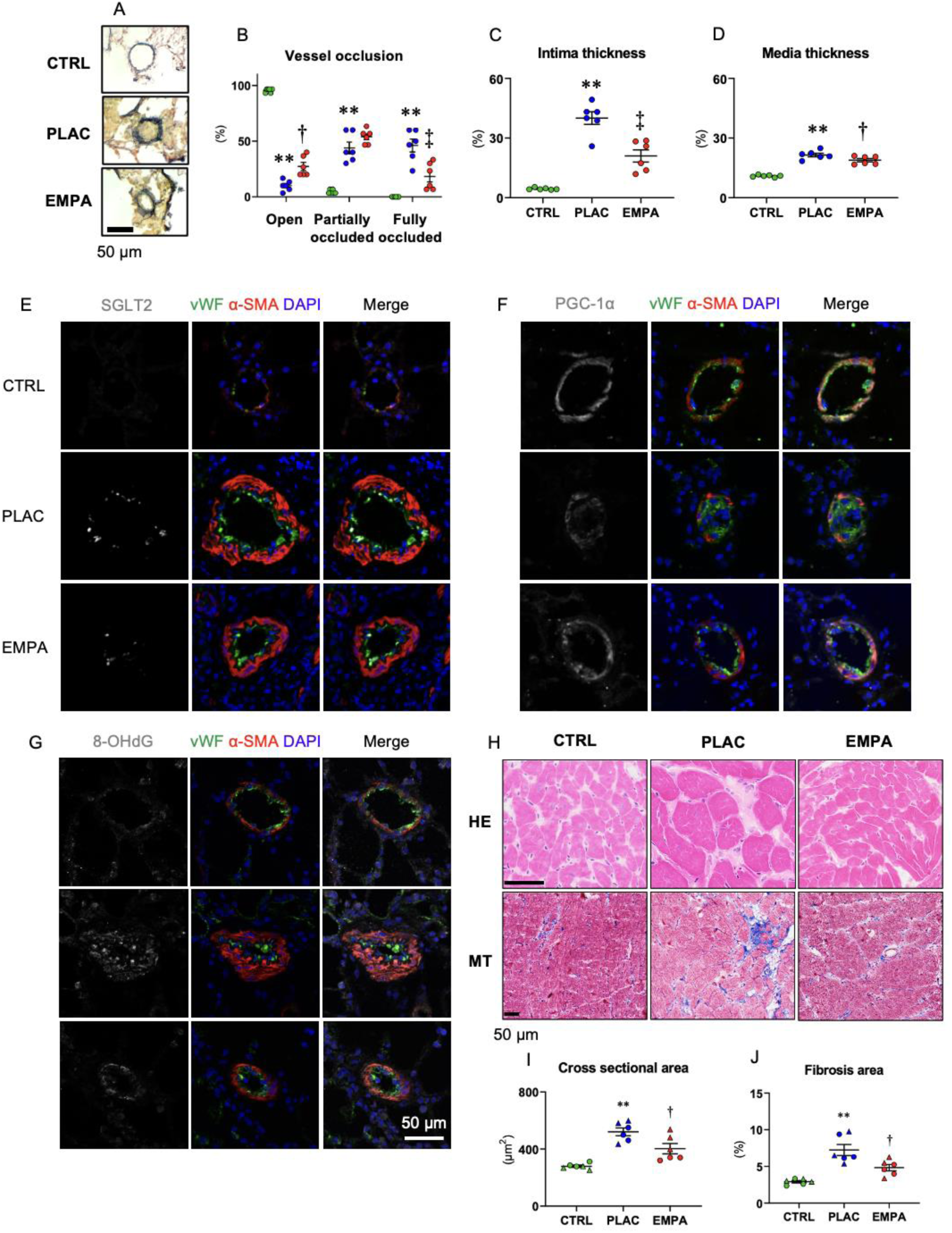
Empagliflozin enhances PGC-1α, attenuates oxidative stress and pulmonary vascular remodeling of PAH rats. **A,** Representative photomicrographs of Elastica van Gieson (EvG) staining from control (CTRL), and placebo (PLAC) or empagliflozin (EMPA) treated PAH rats. **B**, Qualitative analysis of vessel occlusion of PA. Severity of PA occlusion is classified into 3 groups: no evidence of neointimal formation; Open, luminal occlusion less than 50%; Partially occluded, luminal occlusion more than 50%; Fully occluded. We also performed quantitative analysis of intima (**C**) and medial wall thickness (**D**). Representative immunofluorescent staining of SGLT2 (**E**), PGC-1α (**F**) and 8-OHdG (**G**) of the pulmonary artery from control (CTRL), and placebo (PLAC) or empagliflozin (EMPA) treated PAH rats. Section was co-stained with von Willebrand Factor (vWF, green), alfa-smooth muscle actin (α-SMA, red) and DAPI (blue). **H,** Representative photomicrographs of Hematoxylin and Eosin staining (HE) and Masson Trichrome staining (MT) of RV from control (CTRL), and placebo (PLAC) or empagliflozin (EMPA) treated PAH rats. Qualitative analysis of cross-sectional area (**I**) and fibrosis area (**J**). Values are mean ± SEM. Difference was tested by using one-way analysis of variance followed by post-hoc Tukey’s multiple comparisons test. ** p<0.01, * p<0.05 vs. CTRL, ‡ p<0.01, † p<0.05 vs. PLAC.

Representative immunofluorescent staining of SGLT2 (Figure 5E), PGC-1α (Figure 5F) and 8-OHdG (Figure 5G) from rat lung tissue are shown. As can be seen, expression of SGLT2 co-localized with vWF, which was increased in placebo. PGC-1α co-localized with both vWF and α-SMA, and was decreased in placebo and increased in empagliflozin. Moreover, 8-OHdG, a marker of oxidative stress, was increased in placebo and decreased in empagliflozin. Immunoblotting from rat lung homogenates shows that the expression of SGLT2 was increased in placebo, while it was further increased in empagliflozin (Figure S6A and S6B). There was no significant difference in PGC-1α (Figure S6A and S6C).

Figure 5H demonstrates representative HE and MT stainings of the RV. HE staining shows that empagliflozin attenuated hypertrophy of cardiomyocytes (Figure 5I), and MT staining shows that empagliflozin reduced the proportion of fibrosis in the RV (Figure 5J). These data suggest that remodeling of the RV was attenuated by chronic empagliflozin treatment.

### Primary endpoints from the clinical trial

Eight patients with idiopathic or heritable PAH were enrolled to in the trial. Baseline characteristics are presented in Table 1.

**Table 1.** Baseline characteristics of EMPHOWER.

|  | Patients n=8 |
| --- | --- |
| Female – n (%) | 2 (25%) |
| Age, years | 55.2 (13.5) |
| Height, cm | 169.1 (8.0) |
| BMI, kg/m<sup>2</sup> | 29.7 (1.9) |
| BSA, m<sup>2</sup> | 1.96 (0.13) |
| Medical History, comorbidities – n (%) |  |
| Diabetes Mellitus | 2 (25) |
| Atrial fibrillation | 3 (37.5) |
| Hypertension | 2 (25) |
| Smoking | 2 (25) |
| PAH therapy – n (%) |  |
| Monotherapy | 1 (12.5) |
| Double therapy | 6 (75) |
| Triple therapy | 1 (12.5) |

Data are presented as mean (SD) or n (%).
BMI, body mass index; BSA, body surface area; PAH, pulmonary arterial hypertension.

Primary endpoints assessed the safety, tolerability and feasibility of 12 weeks empagliflozin treatment in PAH patients. The first patient was screened on March 1^st^, 2023, and the last patient was included in the study on January 30^th^, 2024, indicating that enrollment was completed in 11 months. Overall, empagliflozin was well tolerated. During the study, 17 adverse events (AEs) were reported, three of which were serious adverse events (SAEs), and one was classified as a serious adverse event of special interest (AESI) (Table S2). All SAEs were related to progression of PAH of which one led to hospital admission and temporal discontinuation of empagliflozin treatment. All SAEs and the AESI occurred in the same patient. In hindsight, this patient may not have been clinically stable at the time of randomization. Disease progression was considered as a natural event in this patient and all SAEs were deemed to be unrelated to the investigational medicinal product (IMP). Consequently, after the patient had stabilized, empagliflozin treatment was resumed and was well tolerated. In total, two patients experienced worsening of PAH during the study. Four adverse events were deemed related to the IMP, all of which were of grade 1 severity.

### Secondary endpoints from the clinical study

Table S3 summarizes the clinical outcomes before and after the treatment with empagliflozin. CMR revealed a significant increase in indexed RV end-diastolic volume (RVEDVi) (from 87 ± 26 to 103 ± 30 mL/m^2^, p=0.037) and RV end-systolic volume (RVESVi) (from 49 ± 21 to 66 ± 28 mL/m^2^, p=0.044). Additionally, there was a significant decrease in RV ejection fraction (RVEF) (from 45 ±10 to 38 ± 12%; p=0.036, Figure 6A), RV global longitudinal strain (GLS) (from −15.2 ± 4.2 to −13.2 ± 4.0%, p=0.002, Figure 6B). No significant change was observed in left ventricular (LV) EDVi, LVESVi, LVEF or LVGLS.

**Figure 6.**
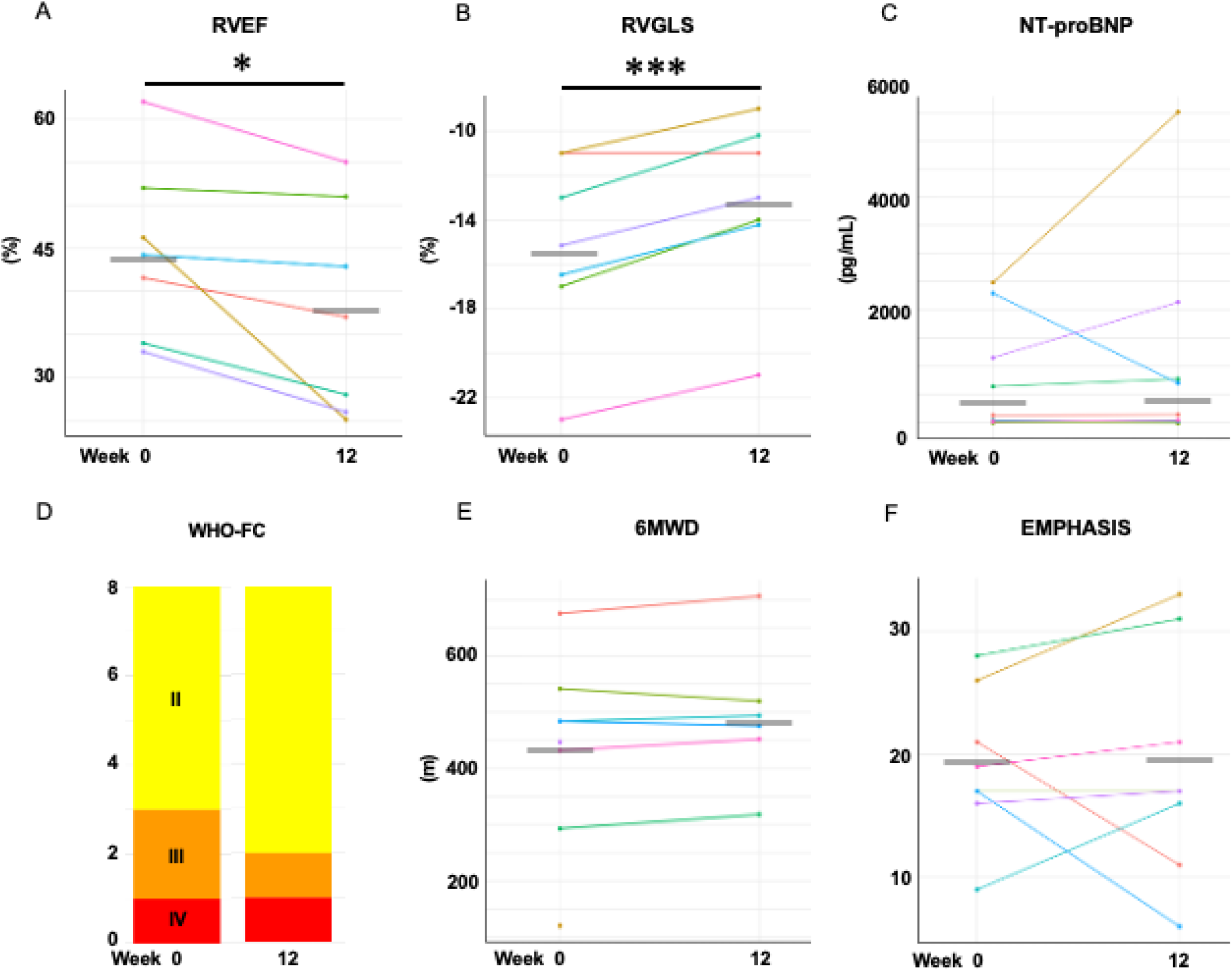
Twelve weeks of empagliflozin did not influence the biomarker, exercise tolerance and symptoms while worsened right ventricular function in patients with idiopathic pulmonary arterial hypertension. Results of right ventricular ejection fraction (RVEF, **A**), right ventricular global longitudinal strain (RVGLS, **B**), N-terminal pro–B-type natriuretic peptide (NT-proBNP, **C**), The World Organization-functional class (WHO-FC, **D**), six-minute walking distance (6MWD, **E**), EMPHASIS questionnaires score (**F)** before and after 12 weeks of EMPA treatment in 8 patients with idiopathic pulmonary arterial hypertension. Difference was tested using a paired sample t-test. * p<0.05, *** p<0.005 vs. Week 0.

We did not observe significant changes in TAPSE, PAAT, TRV, or any of the other examined echocardiographic parameters.

No statistically significant changes were observed in NT-proBNP throughout the study (Figure 6C). An almost 3–fold increase in NT-proBNP was observed in one patient between week 6 and 12, attributable to acute worsening of PAH. The statistical analysis indicated no significant changes in creatinine throughout the follow-up period. All but one patient, who went from WHO functional class (WHO-FC) III to II, remained in the same WHO-FC after 12 weeks of empagliflozin treatment (Figure 6D). The 6MWD did not change over time (Figure 6E). One patient was unable to perform the 6MWD due to an acutely worsening condition.

From baseline to week 12, no statistically significant differences were observed in the EMPHASIS questionnaire scores (Figure 6F). Similarly, across the three domains of the CAMPHOR questionnaire—symptoms, activity, and quality of life, no changes reached statistical significance.

Gene expression of *Pgc1a* and *Bmpr2* were analyzed in peripheral blood mononuclear cells (PBMCs) from 6 patients by qPCR, before and after 12-weeks of treatment with empagliflozin (Table S4). There was a trend of increased expression of *Pgc1a*, however not statistically significant. No significant change was observed in *Bmpr2* expression after the treatment.

## Discussion

In the present study, we demonstrated increased expression of SGLT2 in the intima of remodeled small pulmonary vessels in human PAH. We also showed that increased SGLT2 expression is related to reduced PGC-1α expression, and that by inhibiting SGLT2 in PAH MVECs, PGC-1α as well as several indicators of endothelial dysfunction are normalized. Empagliflozin enhanced the expression of PGC-1α possibly in a SIRT1 dependent way and as such, improved mitochondrial biogenesis and respiration, attenuated oxidative stress and attenuated hyperproliferation of PAH MVECs. In experimental PAH, empagliflozin attenuated pulmonary vascular and RV remodeling, and improved hemodynamics in both male and female rats. In the clinical study, empagliflozin seemed well tolerated with all SAEs in one patient, who had clinical progression that was possibly unrelated to empagliflozin treatment. While on average most functional parameters remained stable, CMR demonstrated worsening of RV function upon empagliflozin treatment.

### Effect of empagliflozin on PAH MVECs

While SGLT2 expression was first described in kidney epithelial cells,^23^ several recent studies reported that SGLT2 is expressed in ECs and that its expression is increased in pathological conditions.^9,24,25^ This study is the first to show the expression of SGLT2 in human pulmonary artery and isolated MVECs. We found that the expression of SGLT2 was increased in PAH MVECs compared to CTRL. Although the exact regulators of SGLT2 are only partially known, our results are consistent with previous reports showing enhanced expression of SGLT2 in ECs in pathological conditions, such as high glucose, hydrogen peroxide,^9^ angiotensin Ⅱ^24^ or palmitic acid^25^ induced endothelial dysfunction.

We demonstrated that empagliflozin enhanced expression of PGC-1α, increased OCR and decreased ECAR, and attenuated oxidative stress and hyperproliferation in PAH MVECs. As mentioned before, PGC-1α is a master regulator of mitochondrial biogenesis,^14^ and has an important protective role in the pulmonary vasculature.^15,16^ To evaluate whether this result is an on-target effect of empagliflozin, we performed knockdown of SGLT2 which also showed enhancement of PGC-1α. Moreover, overexpression of SGLT2 by transduction of plasmid resulted in a reduction of PGC-1α. These results suggest that enhancement of PGC-1α by empagliflozin is not an off-target effect, but results from direct inhibition of SGLT2. Furthermore, alteration of cell metabolism by empagliflozin, demonstrated by increased mitochondrial respiration and suppressed anaerobic metabolism, strongly suggests that empagliflozin improves mitochondrial dysfunction of MVECs.

Takashima et al. reported that luseogliflozin, another SGLT2 inhibitor, enhances expression of PGC-1α and TFAM via phosphorylation of AMPK in cultured brain pericytes.^26^ Other reports suggested that FGF21 is a modulator of PGC-1α.^27,28^ However, we observed no increased AMPK phosphorylation after empagliflozin treatment, and could not detect the expression of FGF21 in MVECs (data not shown). These discrepancies may be explained by cell type differences and/or drug differences. Meanwhile, Umino et al. showed that canagliflozin enhances expression of SIRT1 in porcine renal epithelial cells.^29^ Moreover, another previous report suggested that pharmacological activation of SIRT1 increases activity of PGC-1α.^30^ These results are consistent with our result that enhanced expression of PGC-1α by empagliflozin was, at least partially, induced by SIRT1. We therefore speculate that an SGLT2–SIRT1– PGC-1α axis plays an important role in the pathological changes found in PAH MVECs. Of note, although the mechanism is not known, empagliflozin upregulated BMPR2-Smad1/5/9 signaling in PAH MVECs. This data suggests an additional effect of empagliflozin on pulmonary vasculature in PAH.

Accumulating evidence suggests that SGLT2 inhibitors have multiple means by which they confer endothelial protection.^10,31^ We believe that our findings at least partially revealed this mechanism.

### Effect of empagliflozin on PAH rats

The benefit of empagliflozin in the SuHx rat model may be fully explained by its effects on MVECs. However, we cannot dismiss the possibility that alternative systemic or pleiotropic effects of empagliflozin were responsible for the benefit in this model. As previously reported, empagliflozin is a potential inhibitor of the sodium hydrogen exchanger (NHE), enhances ketone body production and attenuates inflammasome generation via effects on NLR family pyrin domain-containing protein 3 (NLRP3).^6^ Uthman et al. reported that empagliflozin improves viability of cardiomyocytes by inhibiting NHE-1, which consequently reduces cytosolic calcium concentrations and protects mitochondria.^32^ NHE-1 plays an important role in hypoxia induced PH in mice and in PA smooth muscle cells.^33^ Empagliflozin was shown to ameliorate pulmonary vascular remodeling in rats with monocrotaline-induced PH,^11^ a model that lacks profound intimal changes and is highly inflammatory. It is therefore conceivable that empagliflozin exerts an anti-remodeling effect via inhibition of NHE-1 in smooth muscle cells or via an anti-inflammatory effect. Since PAH is a highly inflammatory condition,^34^ and attenuation of pro-inflammatory cytokines may contribute to the regression of pulmonary vascular remodeling,^19,35^ such effects would be highly desirable in PAH patients. Kim et al. demonstrated that empagliflozin suppresses activity of the NLRP3 inflammasome via the production of ketones in diabetes patients with cardiovascular disease.^36^ Although we have not focused on pro-inflammatory cytokines nor on the NLRP3 inflammasome, anti-inflammatory effects of empagliflozin may also be beneficial for PAH patients. Importantly, a recent study from Weis et al. reported that ketone body oxidation increases proliferation of cardiac endothelial cells.^37^ This is an advantage, given the presence of capillary rarefaction in the failing right heart.^38^ However, it could be a disadvantage in hyperproliferative PAH MVECs. Our findings showed that empagliflozin attenuated cell proliferation of PAH MVECs in the *in vitro* condition, independent from ketone body oxidation. In addition, empagliflozin ameliorated intima thickness *in vivo*, probably in the presence of increased urinary ketone concentration. Although it is known that metabolic perturbations play a crucial role in the pulmonary vasculature of PAH patients,^13^ the impact of ketone metabolism on pulmonary vascular remodeling has not been determined.

### Empagliflozin for idiopathic and heritable PAH

Based on the results from our preclinical study, and in light of some unanticipated drug side effects in a previous translational study,^39^ we decided to perform a small PoC feasibility study in PAH patients, with several non-invasive parameters as secondary endpoints. While we observed that empagliflozin was tolerated among this patient group, findings pertaining to right heart function raise some potential concern.

CMR revealed a worsened RVEF and RVGLS after treatment with empagliflozin. Because we did not perform right heart catheterization, we cannot determine with certainty whether RV deterioration was a result of increased mPAP or PVR, or reflected pure RV deterioration. Echocardiography did not show changes in TRV or PAAT. Although the small sample size of this PoC trial prevents us from drawing any conclusions regarding the impact of empagliflozin on RV function, the clear signs of deteriorating RV function cannot be disregarded.

Previous research has shown increased glucose uptake^40^ as well as impaired fatty acid uptake^41^ in the RV from patients with PH. Whether this is an adaptive or maladaptive mechanism remains controversial.^42^ Although the direct impact of empagliflozin on RV function remains unknown, empagliflozin may reduce glucose uptake. In our preclinical study, empagliflozin did not worsen RV function, perhaps as a consequence of a rapid reduction in RV afterload. This effect on the pulmonary vasculature may have been of lesser importance in the PAH patients. Others have shown that neither empagliflozin nor dapagliflozin affected RV function in an RV pressure overloaded model.^43,44^ Clinically, in the setting of heart failure with preserved ejection fraction, dapagliflozin was shown to improve pulsatile pulmonary vascular load and RV function during exercise through the reduction of pulmonary artery wedge pressure (PAWP).^45^ Hemodynamically, in patients with PAH, improvement in RV function cannot be expected through a reduction in PAWP alone, but rather through a decrease in PVR or the direct effects on the RV. An ongoing randomized clinical trial investigates the effect of dapagliflozin in idiopathic PAH and chronic thromboembolic pulmonary hypertension focusing on exercise capacity and hemodynamics (NCT05179356).

### Clinical implication

Empagliflozin has been approved by the U.S. Food and Drug Administration to use in a wide range of clinical conditions, from heart failure to diabetes mellitus, and chronic kidney disease. Although there are several side effects that need to be mentioned, such as urinary tract infection, ketoacidosis and volume depletion particularly in elderly patients,^6^ the safety and feasibility of EMPA for patients with cardiovascular disease^4,5^ and chronic kidney disease^46^ have been well established. Our preclinical findings demonstrated that empagliflozin improved mitochondrial biogenesis and ameliorate pulmonary vascular remodeling in a PAH animal model. Our clinical findings, however, paint a less-clear picture. Although our primary endpoints of feasibility and tolerability were met, there were signs of RV deterioration after 12 weeks of treatment with empagliflozin. Considering the small sample size and open-label design, it is premature to conclude that empagliflozin worsens RV function in PAH. As mixed PAH phenotypes become increasingly common, the use of treatment combinations like empagliflozin in PAH is expected to rise. However, our findings highlight the need for caution when considering such therapies.

### Limitations

There are several limitations of this study that need to be mentioned. First, we used empagliflozin in a concentration of 1 μM to treat MVECs. This concentration was used in several previous reports and a selective inhibition of SGLT2 is expected in this dose range.^10^ Although our knock down data highly suggested that the enhancement of PGC-1α occurred via SGLT2 inhibition, alternative mechanisms are possible. Second, we chose to treat rats with chow containing 300 mg/kg of empagliflozin. Calculating from the consumption of the chow, we assume that ingested between 20 to 30 mg/kg/day of empagliflozin. This is in the dose range used in previous reports.^11,47^ In the clinical study, the dose of empagliflozin was 10 mg/day, which was the same setting for the patients with left sided heart failure, and we have not performed dose adjustment. The patient population enrolled did not reflect demographics seen in typical PAH populations as it was comprised of only 25% women. However, based on the past clinical trials,^4,5^ we speculate that the results were not influenced by sex differences. Additionally, no right heart catheterization was performed and therefore, changes in intrinsic RV function as a result of treatment with empagliflozin could not be determined. The small sample size, absence of a placebo group and open-label nature of the clinical study prevents us from drawing definitive conclusions and may introduce bias, limiting the generalizability and robustness of the findings.

### Conclusion

SGLT2 expression is increased in the PAH endothelium. Empagliflozin treatment attenuates pulmonary vascular remodeling in experimental animal models of PAH, possibly as a consequence of direct effects of empagliflozin on mitochondrial biogenesis and respiration in MVECs. While twelve weeks of empagliflozin treatment is feasible in idiopathic and heritable PAH patients, we did observe signs of RV impairment.

## Disclosure

H.J. Bogaard received research grants and lecture fees from Janssen Pharmaceuticals and MSD. He serves on an advisory board for Novartis. M.L. Handoko received an educational grant from Novartis and Boehringer Ingelheim, and speaker/consultancy fees from Novartis, Boehringer Ingelheim, Vifor Pharma, AstraZeneca, Bayer, MSD, Abbott, all not related to this work. The remaining authors have nothing to disclose.

## Supplemental Material

### Supplemental Methods

Tables S1-4

Figures S1-6

## Data Availability

All data are available at request

## Non-standard Abbreviations and Acronyms

AEs: adverse events
AESI: serious adverse event of special interest
IMP: investigational medicinal product
MVECs: microvascular endothelial cells
PAH: pulmonary arterial hypertension
PGC-1α: peroxisome proliferator-activated receptor gamma coactivator-1α
ROS: reactive oxygen species
RV: right ventricle
SAEs: serious adverse events
SGLT2: sodium glucose cotransporter 2
SIRT1: sirtuin 1
WHO-FC: World Health Organization functional class

